# Care homes, their communities, and resilience in the face of the COVID-19 pandemic: interim findings from a qualitative study

**DOI:** 10.1101/2020.11.10.20229013

**Authors:** Fiona Marshall, Adam L Gordon, John RF Gladman, Simon Bishop

**Author notes:** Corresponding author details: Fiona Marshall, Division of Medical Sciences and Graduate Entry Medicine, Derby Medical School, Royal Derby Hospital. Derby. DE22 3NE.

## Abstract

**Background:** From late February 2020, English care homes rapidly adapted their practices in response to the COVID-19 pandemic. In addition to accommodating new guidelines and policies, staff had to adjust to rapid reconfiguration of services external to the home that they would normally depend upon for support. This study examined the complex interdependencies of support as staff responded to COVID-19. The aim was to inform more effective responses to the ongoing pandemic, and to improve understanding of how to work with care home staff and organisations after the pandemic has passed.

**Methods:** Ten managers of registered care homes in the East Midlands of England were interviewed by videoconference or phone about their experiences of the crisis from a structured organisational perspective. Analysis used an adapted organisational framework analysis approach with a focus on social ties and interdependencies between organisations and individuals.

**Results:** Three key groups of interdependencies were identified: care processes and practice; resources; and governance. Care home staff had to deliver care in innovative ways, making high stakes decisions in circumstances defined by: fluid ties to organisations outside the care home; multiple, sometimes conflicting, sources of expertise and information; and a sense of deprioritisation by authorities. Organisational responses to the pandemic by central government resulted in resource constraints and additional work, and sometimes impaired the ability of staff and managers to make decisions. Local communities, including businesses, third-sector organisations and individuals, were key in helping care homes overcome challenges. Care homes, rather than competing, were found to work together to provide mutual support. Resilience in the system was a consequence of dedicated and resourceful staff using existing local networks, or forging new ones, to overcome barriers to care.

**Conclusions:** This study identified how interdependency between care home organisations, the surrounding community, and key statutory and non-statutory organisations beyond their locality, shaped decision making and care delivery during the pandemic. Recognising these interdependencies, and the expertise shown by care home managers and staff as they navigate them, is key to providing effective healthcare in care homes as the pandemic progresses, and as the sector recovers afterwards.

## Introduction

COVID-19 produced a significant crisis in English care homes(1). Care home residents are especially at risk of COVID-19 because of their age, co-morbidities, prevalent frailty, cognitive impairment, and functional dependency(2). Residents are also rendered more vulnerable by virtue of frequent close and personal contact with other residents, and carers who attend to their needs. COVID-19 spread rapidly through care homes and, despite the efforts of staff, many homes were devastated by large numbers of deaths and associated illness in their residents and staff. In the year to 19 June 2020, there had been more than 30,500 excess deaths among care home residents(3), and by 26 June 2020, there had 268 deaths involving COVID-19 among social care workers(4).

When COVID-19 first arrived in the UK, Public Health England (PHE) announced that care homes were not at high risk of exposure(5). A UK national lockdown was announced on the 23^rd^ March 2020(6). Although care homes were referenced in early guidance about hospital discharge on 19^th^ March 2020, the first government guidelines specifically focussed on care homes were not produced until 2^nd^ April 2020(7). Recommendations around infection control, testing, workforce mobility and hospital transfers in care homes underwent multiple changes during the period from February to July(1). At the same time, as part of infection control, organisations which routinely provide support to care homes, including general practitioners and community based multidisciplinary teams, were asked to minimise contacts with care homes to those deemed essential. This meant rapid reconfiguration of the support services that care home staff routinely rely on for advice and guidance.

Despite gaps in the public policy response, care homes were not equally affected by COVID-19 during this first wave of the pandemic. For example, over half of COVID-19 deaths in Scottish care homes during the first wave of the pandemic came from outbreaks in only 13 homes, and similar epidemiology was seen in the other UK nations(8). Some of the differences in experience between homes will have been due to chance and external forces outside the control of care home staff, but outcomes will also have been shaped by how staff navigated this complex and rapidly changing environment to make critical decisions about care delivery.

Our research explores the experiences of care home managers as they navigated the challenges raised by COVID-19. These experiences took place against a background of longer standing challenges affecting long-term care in the UK. Care homes are critical to health and social care provision because they provide care for people who are vulnerable, frail, often approaching the end-of-life and no longer able to receive care in other settings(9). There are 15,000 care homes in England, providing approximately 430,000 beds. 50% of care homes are run by profit-making business, 47% are not for profit and 3% are state run(10). Most care home staff are middle-aged or older and come from a black and ethnic minority background(11). Care home work is often regarded as being of low status and many working in the sector only receive minimum wage. Despite several independent commissions and recommendations for reform(12,13), it is widely recognised that the long-term care sector in the UK is beset by systemic challenges. These include a lack of funding, fragmentation of supply leading to large variations in the quality and quantity of care, poor integration with the wider health and social care system, and problems in staff recruitment, retention and training(13,14). This is now recognised in the Ageing Well components of the NHS England Long-term plan(15). This includes specific objectives around Enhanced Health in Care Homes, but these are yet to be implemented(16). The increasing dependency of the care home population has led to role-extension for care home staff, who now attend to aspects of care that would previously have been regarded as the responsibility of the National Health Service (NHS)(17). This has not been accompanied by commensurate re-allocation of funding and this has led to inequity of resource provision, with expenditure on care homes in the UK well below the (OECD) average(13).

Given the above, we aimed to examine care home managers’ responses to the pandemic with consideration given to their location in a complex health and social care system. We set out to learn lessons that might inform more effective approaches to working the care homes as the pandemic progressed, and which might prove useful beyond the pandemic as the sector recovered. This work is part of a larger ongoing project focussing on how care home managers connect and interact with healthcare services and providers, but we have chosen to present this interim analysis given the urgency of the current crisis.

## Method

To provide a framework for understanding the challenges faced by care home managers, we adopted an organisational systems approach. This views organisations as involving complex and contingent relationships between dynamic elements, rather than being static rational-bureaucratic entities. In this instance we focused particularly on ‘interdependencies’ within homes, and between homes and external organisations. Interdependencies are defined as “a relationship between two or more elements (e.g. roles, units, work processes) that are linked or mutually reliant on each other.”(18) Interdependencies within organisations are often seen to be important because they provide insights into where there may be tension, collaboration, or ambivalence within organisational systems. This approach was chosen following initial conversation with participants, which suggested that their experience of the pandemic, and their ability to respond to it, was closely ‘tied in’ to the relationships and processes of the wider health and social care system. In other words, while care homes are often felt to be left out of mainstream health and social care policy in the UK, in the face of the pandemic, the web of mutually dependent relationships was brought to the fore.

From this initial insight, and following an inductive-deductive approach, we considered a series of interdependencies, interfaces and points of criticality and uncertainty likely to have a bearing on care home managers’ work. Starting from the work of Worren(18), we focussed on care processes and practices, resources and governance. We developed these ideas by rapid thematic analysis of the data as each interview was conducted. Through this process, newly formed networks emerged as being of critical importance and so we focussed particular attention on these in later interviews and transcripts.

Ten registered care home managers were recruited from within the East Midlands of England. All were from homes registered for care of older people. Each manager had been employed at the home for at least six months prior to the interview. Participants were identified through existing care home networks, established by the research team through previous work conducted in and with the sector. We used these as a starting point for a snowballing approach to recruitment, asking members who had previously participated in research to help us identify care home managers who had not been engaged in research before. We supplemented this by asking professional care and research networks to publicise the study using social media and electronic mailing lists. We chose to focus on managers new to research because many care home managers that we had routinely interacted with in the past through our research had engaged with online support forums alongside senior clinicians and academics(19). We considered that such experiences were likely to be atypical and we wished to explore broader accounts of how care homes responded to the pandemic.

Consent was obtained by email. Interviews were semi-structured, remote and conducted using either telephone or videoconferencing software. They lasted on average 35 minutes. Each interview was taped and transcribed verbatim by a qualified transcriber.

## Results

The results focus on the relationships and changing nature of the key organisational ties as described by the practitioners and leaders as they experienced the pandemic. They also evidence the nature of interdependencies and the ways in which care home staff commonly experienced issues within each of these interdependencies.

We present three main interdependencies: care processes and practices; resources; and governance. These issues were often intertwined but we present them here individually for clarity of explanation and understanding. Within each of these three interdependencies we highlight the problems and sources of resilience.

### Interdependencies of care processes and practices

The first form of interdependencies were in the care processes and practices of the care homes, strongly shaped by their position as the endpoint of many NHS ‘care pathways’. As the pandemic broke, managers were faced with the challenge of delivering a broader range of (new) care duties as the wider health and social care system changed around them. This included taking on care previously delivered by trained clinicians within primary and community care. Since many GP practices ceased visiting during the pandemic these regular care duties, such as injections and wound care, fell to the care home staff. Adapting to the restrictions of lockdown required new approaches to care, managing everyday care demands alongside COVID-19 requirements. This included developing approaches that enabled staff to simultaneously care for those without COVID, those who were COVID positive, and those recovering post-COVID who needed rehabilitation. There were additional demands of new COVID-related cleaning and administrative activities, for example, deep cleaning after COVID-infection and completing mandatory online reporting templates for the NHS.

Daily routines also changed for residents. In some homes, residents were encouraged to participate in meaningful activities as an integral part of their dementia care. Residents assisted with infection control by taking on roles as door handle cleaners and general helpers. Staff were aware of the impact of the changes in routine, especially among residents with dementia and without any family networks. For these residents, the care home staff could not replace embedded routines, established before the pandemic, because of the increased demands on their time. The sudden cessation of routines led to expressions of anger towards the staff:

> ***“It’s heart-breaking, to be honest you know, I don’t want to be in their shoes. … we are trying our best but it’s not the same you know. I mean one particular lady I’m thinking of, our owner used to take her every Monday to the cake shop and they used to go for a coffee and have a cake, always had blueberry muffin and bring something back for the staff. And she couldn’t understand why she couldn’t … I mean she could see the minibus parked there and she was like getting really angry. Why can’t I go out you know, why can’t I go for a drive?” CH8, July 2020***

The increased death rate associated with COVID-19 in some homes caused a number of issues. In addition to the severe emotional toll of coping with dying patients, managers were asked, in places, to verify death without what they regarded as adequate training to do so. Funerals could not be attended by staff, family or residents, which was seen as adding to the emotional pain of death. While managers spoke of the ways in which digital media enabled residents to “attend” services where possible, this was often difficult for those with dementia. The idea of memorial events or services after the pandemic was regarded as being of spiritual and psychological importance by several respondents. Notably, none of the participants mentioned the absence of family members during the dying process, perhaps an indication of how lockdown had become normalised by the time of the data collection. One manager spoke of her devastation at accepting a previously unknown resident as a new admission to the home, only for the virus to spread through the home with the loss of 7 residents and 1 member of staff to COVID-19:

> ***“I blame myself for*** <u>***every***</u> ***death. I didn’t turn them away. A 96 year old in the back of the ambulance at 11pm at night. They knew we had a bed. But we only had a bed in the green zone. I could only use the green zone. Two days later their test came back positive. Too late then”. CH5 July 2020***

In light of these experiences, managers reported feeling isolated, as if they had been left to deal with the rapidly increasing mortality and multiple demands of the crisis alone. This feeling was exacerbated during the early phases of the pandemic by the fact that there was little specific care home practice guidance to draw upon except pre-COVID legal and practice guidance. A commonly cited dilemma was where a resident hospitalised for COVID-19, could best be cared for by rapid return to their care home, even whilst they remained infective. Such decisions required daily emotional, moral and logistical energy as the managers struggled to make sense of the, sometimes conflicting, needs of individual residents and the need to protect the other people resident in the care home.

Managers reported that the skills and determination of care home staff was a source of resilience. Managers spoke of pride in their teams, who worked with tenacity and creativity, and recognised skills that had previously not been required. For example, some staff proved adept at building relations with the local community, managing to secure supplies from shopkeepers when formal supply chains were unreliable. Managers identified, and became more confident, in their leadership abilities as they led these teams.

> ***“I always thought I was just thinking on my feet, nothing more. But you know, this has shown me that I am far more. I can bring together my flock. I can lead. I show them every time that I mean what I say and do. They all know infection control inside out. They are all so great at it. Even my mobile foraging residents do infection control because they are good at it. They have their own cleaning kits. Bring them all on board***.***” CH3 June 2020***

Two homes deployed staff who spoke fluent Italian and Cantonese to monitor international media stations early in the pandemic. This enabled them to gauge the seriousness of the situation daily and was cited as key to the decision, in these two homes, to lockdown by the 2^nd^ March 2020, three weeks earlier than the national lockdown and in direct opposition to all English government guidance at that time. This included a refusal to accept any admissions, family visits and, at one home, staff moved into the home for periods of 6 weeks continuously in rotation. Both homes (independently) drew on international sources of infection guidance in advance of any UK guidelines:

> ***“we knew this wasn’t a flu thing early. We had a member of our core staff and she gave daily updates to our management from Italian News. Old people were suffering. The Italians love their old folks despite what our media was saying. So we decided as our residents are our folks. We love our residents. So we locked down”. CH4 June 2020***

In addition to the ‘hidden’ skills of staff, some managers reported consciously adapting their leadership style towards a more hierarchical “military” style of command. Managers were required to make rapid decisions on practice as well as provide reassurance to their staff, many of whom were frightened and exhausted. Several managers devised “COVID-19 command centres” which met daily to review emerging evidence and data, and devise new care practices. These responsive management structures enabled managers to redeploy care home resources in novel ways in response to the pandemic.

> ***“The one thing we have started is we’ve got a big minibus, so we’ve got a few people who drive it and they are ringing relatives up and we’re doing like a drive-by wave or we’ll stop and they can talk through the window with them and things like that. Just to get the residents out of the home but they’re only on a bus***.***”CH4 July 2020***

Some managers reported closer relationships with their residents, despite physical distancing, because of the frequency and intensity of time spent together. Some managers reported improved health outcomes among their residents, with fewer infections, improved weight control and mobility. Some homes commenced new routines to help residents’ cope with lockdown, including daily exercise programmes, healthy eating regimes and daily activities. Large scale gardening and decorating projects had been undertaken with the residents as co-producers. These innovative approaches went some way to compensate for the lack of visiting from families and external agencies that would usually help with entertainment and occupation.

> ***“they’ve done really well. We remind and explain most days why the masks. They help us out. For the most confused and mobile we have given a cleaning carrier, so they can take part and we know where they are. We can’t isolate them. Washing hands is by washing up a few cups. We normalise it all…” CH2 June 2020***

## Interdependencies of resources

The second form of interdependence was in the supply networks for key resources needed to cope with the pandemic. One of the main issues was provision of personal protective equipment (PPE). During the early phases of the pandemic, this was in short supply to the care home sector with an increase in general demand and priority given to the NHS due to national shortage. In addition to core PPE requirements of aprons, face masks and gloves, some care home residents presented with symptoms such as diarrhoea, which demanded additional supplies of incontinence pads and extra laundry. These extra resource demands rapidly utilised the standard stocks of PPE held by most homes as a requirement for any infectious outbreak.

> ***“Overnight we lost our supplier of over 30 years to the NHS. So suddenly no PPE, no aprons, no gloves, no pads for the diarrhoea. They don’t mention the diarrhoea. So we had to pay 4000% more. One bottle which used to cost us £2***.***50 is now £52. So do the maths. We are a charity. We had to use soap and beg…the NHS took all our supplies” CH9 July 2020***

As care homes became more formally incorporated in NHS supply chains, care homes continued to struggle to obtain adequate supplies. Deliveries were unpredictable and either smaller than anticipated or contained equipment of uncertain quality. Care home managers who worked for organisations with longstanding relationships with suppliers were particularly angered about this as they felt this was a forced dependency for equipment which they had already procured prior to, or early in, the pandemic.

> ***“the issue with that was when the Government announced they would provide PPE equipment free of charge to all care homes and surgeries and whatever, which was fine but what they were doing was they were intercepting stock that we would have normally purchased from our suppliers. So then our supplier was saying we can’t send it out because we’ve got to give it to the Government. And so our options for buying stuff then was quite limited” CH6 July 2020***

Managers also reported financial difficulties in the face of extra running costs for staffing required to cover sick leave, topping up wages to supplement the government furlough scheme for non-essential staff, and paying staff during shielding or self-isolation due to quarantine. Most home owners in our sample provided staff with bonus payments and free meals in recognition of the extraordinary commitment shown during the pandemic, but this added to financial strain. None of the homes operated zero-hour contracts or cut sick pay to the bare statutory minimum.

Staff resource, meanwhile, was stretched. Staff worked hard to cover rota gaps. In some instances managers refused to accept agency staff that worked across multiple sites, recognising that they might be vectors of spread for infection. This meant that staff had to work in ways that would not be tolerated in care home before, or in larger organisations during, the pandemic.

> ***“that poor lass in that nursing home, it was the registered nurse, she did three shifts back-to-back and ended up staying overnight. CH8 July 2020***

Space and environment placed important constraints on homes’ ability to respond to the pandemic in the way that was being recommended in government documents and external guidance. The concept of setting up cohorted areas or “red zones”, was challenged both by the amount of appropriate space in buildings that were not designed primarily as healthcare facilities, and by the availability of staff, and patient care resources. Additional beds, bathing equipment, hoists and other lifting equipment were required depending on the residents being moved to these areas. Making safe decisions about infection control in the context of environmental constraints forced by buildings was seen as very demanding by managers.

Home managers reported some sources of resilience to help combat these resource shortages. First, informal networking between care homes and domiciliary care agencies who were recipients of PPE, enabled diversion of resources to homes where the quality of PPE was inadequate in quantity or quality. Some homes used donated bedding, produced their own PPE from duvet coverings, or mobilised local businesses to manufacture items. Respondents also reported general practitioners, veterinarians, dentists, and heavy industry arranging to donate and deliver boxes of PPE urgently to the homes after news of shortages was reported in local and national media. Ski googles, sanitiser gels, bleach, toilet rolls, protective ventilation masks from heavy industry, home-made coveralls and extra bedding were also delivered to homes.

> ***“… a community school, …they made us all face shields. And they were all free, we got a hundred free. So we took what we needed and then gave the rest to all the supported living providers who actually were not really given any PPE because they weren’t entitled to it but they do … you know we’ve made sure that they’ve got them” CH4 July 2020***

In addition, informal networks of companies and voluntary organisations from the immediate locality also supplied other resources such as tablet computers, gardening and arts equipment. Tablet computers were particularly useful because they enabled residents and staff to communicate with others such as family via video-calling software. Numerous local organisations reached out *towards* the care homes with offers of practical help, knowledge sharing and financial support. We identified contributions by local businesses, schools and local third sector organisations of taxi rides, activity materials, bedding and food for staff, car maintenance, garden shelters to facilitate family visits, carpentry to zone buildings and singing/exercise/dancing outdoors for residents to enjoy watching. The care homes continued to value the deliveries of dementia friendly sleeves and “twiddle muffs” from knitting groups and gestures such as potted plants. These seemingly small items helped to make the teams feel part of their community.

In addition to physical goods, managers reported an increase in volunteering of time and expertise from the local community. Local hospices volunteered their expertise in managing death and bereavement among staff and residents, with some hospice staff visiting the homes to offer direct end-of-life care to residents and to support staff. Online counselling and courses, personal phone calls and videoconference follow-ups were offered with specific support for the manager. As well as this, managers especially valued the ongoing support of families and local schools who regularly provided letters and artwork for the homes especially where staff were living in as part of the lockdown.

> ***“We’ve actually had the school, the local school sent us pictures from the children, chocolates for the residents, they’ve been absolutely lovely” CH4 July 2020***

Networks with other regional health providers were also crucial in dealing with clinical resource and knowledge issues. Pharmacists and hospices agreed to collectively store and maintain additional supplies of end-of-life medications and syringe drivers for use by several homes to facilitate rapid delivery as required and avoid delays in care. Care homes put these arrangements in place well ahead of changes to the law in May 2020 which enabled care homes to hold stocks of end-of-life medication(20). These “work arounds” for national protocols proved to be pragmatic and responsive, and ensured end-of-life medications were always available to the care homes. Larger groups of care homes, who had more resources to stay updated with changing government guidelines and evidence also reported sharing their knowledge with independent homes. New networks were being created despite previous barriers of geography and the intrinsic competition between providers which had previously limited links between homes.

## Interdependencies of governance

The final form of interdependence that affected the impact of the pandemic on care homes related to governance. While there is considerable variation across the UK in the governance of homes depending on, for example, their size and ownership structure, what was noticeable was that there was considerable change in governance over the course of the pandemic. In particular, home managers reported governance systems and structures becoming fractured with a perception that there were points of inequities and a lack of fairness in the application of rules, guidance and available support across the health and social care sector. Within this, care homes were often subject to wider systems of governance, enforced by state agencies, without necessarily benefiting from them.

Care home managers felt that they were held accountable by regulatory agencies for the safety of their residents, while also being expected to follow general guidelines inappropriate for their settings. This challenged their accountability. For example, in an instance in which a care home ‘locked down’ earlier than national guidance to protect residents, complaints were made by families to the Care Quality Commission (CQC), which regulates and licences English care homes, about abuse of civil liberties. Difficult interactions with the CQC ensued, which the manager felt questioned her judgement, even though such approaches were adopted nationally a few weeks later.

Reporting requirements imposed by external agencies multiplied quickly. Managers described the need to duplicate information inputs to meet the requirement by multiple external organisations, with no tangible benefit to the care home. A further concern for care home managers was that rules being imposed, with regard to care homes, were interpreted and managed differently by staff within and outside care homes. This was particularly evident during transfers of care, which were points of high risk of infection, and for which a number of guidelines had been developed. There was a sense that hospital staff and care homes staff either interpreted, or adhered to, these differently. This led to conflict and stress.

> ***“And it’s the constant debates with discharge coordinators and ward sisters is you know, really poor because they’re not following what we’re getting told. Now whether it’s their guidelines or totally separate but it still states in the guidelines for NHS discharges that they’ve to swab them but they’re not doing it. Community hospitals are doing it, they’ve done it, but the acute hospitals aren’t***.***” CH2 June 2020***

Resilience in the face of changing governance requirements, was largely felt to come from the strength of existing ‘bottom up’ social networks. As one example, the following describes the local relationship between a care home manager and GP who sought to close a planning gap in the pandemic response by working collaboratively with other homes to evidencing issues around short staffing, forcing regional planners to develop contingency plans.

> ***“…So we kind of spent the weekend going through escalating and it got highlighted within the CCG and the care home cell you know, that…***.***people [couldn’t] fulfil their registration of their organisation if they’d not got staff to keep it open?***… ***there was not really a thought for how we were going to … what continuity we’d got with the community…plans were put in place quickly afterwards***.***” CH8, July 2020***

Previously firmly established ties with adult social workers and specialist NHS primary care staff remained at least as strong during the pandemic. Some homes also reported more supportive ties with individuals in regulatory organisations such as the CQC, that they had previously not trusted. Third sector contacts, including in hospices, also proved invaluable, and were seen to act as knowledge brokers and boundary spanners between smaller, isolated care homes, acute NHS Trusts and Local Authorities.

> ***“…they’ve been brilliant the hospice. And they’re still fighting … and they’re fighting for us as well to get the testing, for all the staff and the residents on a regular basis” CH3 July 2020***

Groups of care home staff collectively shared information such as policy changes via closed forums using online social networking technology. While this was seen to assist in the spread of relevant governance information, it should be noted that, for some staff, these were overwhelming in the volume messages and updates. Wearing PPE meant that mobile phones could not be accessed for long periods of the shifts and it was stressful to trawl through accumulated messages in breaks or at the end of shifts. Two participants admitted leaving these groups because of fatigue and “COVID-19 guilt”, which came about when having to talk about cases in their homes through peer networks.

## Discussion

The main findings of this research are the interdependencies between care homes, and external organisations and individuals, relating to care processes and practices, resources and governance. Centrally organised governmental responses to the pandemic often had counterproductive effects in care homes, resulting in resource constraints and additional work, and impairing the ability of staff and managers to make decisions about care of their residents. Much of the resilience in the system was a consequence of dedicated and resourceful staff using existing local networks, or forging new ones, to work around the challenges brought forward both by the pandemic and the central policy decisions intended to deal with it.

Some of the findings in this paper describe a care home sector placed in considerable jeopardy both by the pandemic and organised responses to it. The latter were insufficiently cognisant of, and insufficiently expert in, how care is delivered in and by care homes. A countervailing narrative comes from the way in which local communities rallied around care homes to help them through. There is evidence, at times, that care homes were inadequately considered and valued by those making and delivering policy. There is also evidence of largely unconcerted support efforts by members of the community, who clearly valued care homes, their staff, and the work that they do. Important lessons emerge, both for the remainder of the pandemic, and how care home staff are supported and facilitated in their role to care for their residents in the longer term.

Previous studies have shown how care home managers and staff play an essential role in brokering relationships between residents and relatives, and statutory health and social care providers. This has been shown to be the case for urgent and planned healthcare(21,22), in end-of-life care(23) and when working to improve and develop services(24). This study adds to this by showing the essential leadership and brokering role that managers and staff played in maintaining continuity of care during the early stages of the COVID-19 pandemic. Many of the strategies adopted by managers, relying largely on their own experience and intuition in the absence of coherent national guidance, reflected those which were reported to be effective in controlling the pandemic in care homes during the initial COVID-19 epidemic in Wuhan Province, China(25). These included establishing clear leadership hierarchies, regularly reviewing policies in light of emerging evidence, developing organisation-specific infection control policies, working to establish reliable supply chains in the face of shortages, and working with residents to maximise activity and engagement. It was care home managers, working with local communities, that ensured these were in place, not central government policy.

Our findings show that healthcare and care homes are interdependent, and that these interdependencies are complex. Our findings show that when central and local government increased the formal reporting requirements placed on care homes, ostensibly to better understand their needs, it resulted in duplication of effort for an already overstretched staff, and risked jeopardising resident care. When PPE supplies were centralised, it robbed those care home managers who were usually very competent managers of their supply chain, of control over their pandemic response. Multiple previous studies have reported the need to engage care home organisations, managers and staff in changes to the design and configuration of care in the sector(17,26,27). The findings presented in this paper show the potentially serious ramifications of the failure to do so. A valuable finding was the way in which are homes are interdependent upon, and embedded within, their local communities. When formal healthcare and social care were slow to provide the necessary support to care homes, third sector organisations, schools, shops, manufacturers and hospices stepped in.

Care homes have historically been held, within the English system, as being private providers in direct competition with each other. Recently there has been evidence that care homes may be prepared to work in a more concerted way as a sector when they engage with research networks(28) and in regional improvement programmes(29). The findings presented here build on those from other studies conducted during the pandemic(19) which showed have shown the preparedness of care home providers to work together not only to provide peer-support, but also advice and expertise, and even critical supplies.

This study has some limitations. Only care home managers were interviewed and not representatives of the NHS, public health organisations or social care. It is only a small sample (n=10). The majority of the study was conducted by one ethnographer and so may only present one worldview. The study continues to recruit and so these findings are interim ones based on ten care homes, all located within the East Midlands of England. There may be regional variation that these results fail to capture.

These findings have important implications for how statutory health and social care providers work with care homes during the second and subsequent waves of the pandemic and beyond. They also provide useful insights as improvement work with care homes continues beyond the pandemic and as national government proposes reconfiguration of the care home sector. It is important to recognise the ability of care home staff to identify and solve emerging issues in care homes. Care homes are under-represented at all levels of decision making – this must be rectified. The importance of care homes to their local communities, and the preparedness of communities to support care homes, is an important new finding. This means that stakeholders in decisions about care home provision are potentially wider than previously recognised. This requires that consultation must be broader when changes are implemented, but it also provides potentially novel opportunities for how care home staff can be supported by communities. Finally, the care home sector has the potential to act in a more concerted, and collaborative way, than has been historically recognised by NHS and social care providers. This potential for these networks to provide peer support and logistical solutions is an important future area of enquiry.

## Ethical Approval

This study was approved by the Faculty of Medicine & Health Sciences Research Ethics Committee, University of Nottingham-02-0420.

## Data Availability

This work presents an interim analysis of interview transcripts that comprise part of an ongoing research project. No data are publicly available at present.

